# Autistic Adults’ Perceptions of the Adequacy of their Supports and Disability-Related Information: Instrument Validation & Preliminary Correlates

**DOI:** 10.64898/2026.10.19.26365938

**Authors:** Rachel K. Schuck, Ian Moura, Mary J. Baker-Ericzen, Liu-Qin Yang, Katherine McDonald, Steven Kapp, Mirko Uljarevic, Julia Love, Joseph Vera, Willi Horner-Johnson, Rachel Kripke-Ludwig, Zack Siddeek, Christina Nicolaidis

## Abstract

Autistic adults face considerable challenges in receiving adequate support, from both paid/formal (such as from disability services) and unpaid/informal (for example, from friends and family) sources. Having adequate support can lead to better health and well-being outcomes. However, most instruments designed to assess adequacy of support were not made specifically for/with autistic adults. This paper presents a psychometric validation of the AASPIRE Adequacy of Supports and Information Scale (AASIS), a community-informed adaptation of a previously developed instrument (the Quality of Care and Support scale; Lucas-Carrasco et al., 2011), designed to be relevant and accessible to autistic adults. We also used the AASIS to explore correlates of perceptions of support in a sample of 798 autistic adults. Our confirmatory factor analysis supported a two-factor structure for the AASIS (factor one: Supports; factor two: Information). Further testing confirmed the instrument was invariant across several groups, including race/ethnicity, gender, intellectual disability, communication fluency, and self-versus proxy-report. Regression analyses suggested that the way in which participants joined the study (i.e., through community advertisements or via established healthcare/disability services) impacted whether they viewed their support as adequate. Additionally, a higher number of adverse childhood events and greater experience of discrimination were related to worse perceptions of support adequacy across both factors. Our findings support the preliminary validity of the AASIS across a variety of autistic people, though further testing is warranted. Findings also point to the importance of addressing the needs of autistic people who are minoritized and/or who have complex trauma histories.

## Introduction

Having access to and receiving high-quality support - that is, support that is accessible, appropriate to one’s needs, and responsive to individual preferences (Burke et al., 2024) - is an important aspect of quality of life for autistic individuals (McConachie et al., 2020; Renty & Roeyers, 2006). Previous researchers highlight support as an important topic for future research due to its connection to wellness for autistic people (Gotham et al., 2015; Nicolaidis et al., 2025). However, researchers have not developed existing tools to assess adequacy of disability-related support with input from the autistic community or validated them for use with this population. This can lead to issues with the accessibility, clarity, and validity of such instruments (Nicolaidis et al., 2020). This paper addresses this gap by presenting the psychometric validation of an instrument to measure the AASPIRE Adequacy of Support and Information Scale (AASIS), a tool for measuring self-reported adequacy of support that was adapted in collaboration with and validated in a sample of autistic adults.

Autistic adults often receive support from a variety of individuals and organizations, such as family members, friends, healthcare facilities, and disability services. Like other people with disabilities, autistic adults may receive either paid/formal support, unpaid/informal support, or a combination of the two. Formal support is generally provided by organizations that employ professionals for this purpose (e.g., doctors, therapists, residential care staff), paid for either directly by recipients and their families or through governmental programs, and distributed in accordance with the laws and regulations that govern a specific area (Lu et al., 2020). Conversely, unpaid/informal support, sometimes referred to as “natural support,” is provided by non-professionals with a relationship to the individual such as family members, friends, and neighbors, often in ways that arise organically (Friedman, 2025). Support may be blended, and in some cases, autistic adults are able to participate in programs that allow them to hire family members, friends, or other individuals they may choose to deliver formal support, adding to the complexity of differentiating between paid and unpaid or formal and informal support (Friedman, 2025). Support can also vary greatly in purpose and method. For example, support could include an occupational therapist teaching daily living skills, someone listening to a friend in distress, or a Facebook support group member providing recommendations on quality service providers in the area.

Regardless of whether support is formal/paid or informal/unpaid, research shows that it is important for the health and wellbeing of people with disabilities, including autistic adults. Previous studies demonstrate that disabled people who receive greater amounts of support like attendant care, homemaking services, and home-delivered meals have a lower risk of hospital admission (Xu et al., 2010). People with intellectual and developmental disabilities (IDD) who have formal organizational support in place are more likely to access routine and preventative healthcare and have better health outcomes than those who do not (Friedman et al., 2020), and autistic adults who receive a greater number of person-centered supports report more positive quality of life outcomes than those who receive fewer supports (Friedman, 2025). People with disabilities who receive unpaid/informal support from natural support networks such as family, friends, and neighbors are more likely to be in the best possible health, achieve personal goals, live in integrated environments, and be free from abuse and neglect (Friedman, 2025), and autistic people (Renty & Roeyers, 2006) and people with IDD (Friedman, 2021) who receive unpaid/informal support have greater quality of life than those who do not.

While support can play an important role in promoting health and wellbeing for autistic adults, these benefits are only possible when the support autistic adults receive is adequate and responsive to their needs. Assessing the adequacy of support and care received by disabled individuals, including autistic people, is thus important from both clinical and research standpoints, and these assessments can serve multiple purposes. First, adequacy of support can be conceptualized as an outcome measure, with service agencies using clients’ perceptions regarding their care as an indicator of how well the agencies are providing those services.

Second, adequacy of support assessments can be used by professionals to better tailor services to individuals. By understanding client perceptions regarding their care, professionals can pinpoint areas of improvement. Assessment of adequacy of support can also be used in research to uncover organizational and individual factors that predict satisfaction. This can further identify areas that can be targeted to improve perceived quality of support, which may facilitate the health and wellbeing of autistic adults.

To date, research on support for autistic adults has mostly relied on parent/caregiver reports and has primarily focused on the lack of appropriate services. This suggests that adequacy of support is a major concern and that there is a need for continuum-based scales that capture a total absence of needed support as the lowest level of adequacy. A recent parent interview study of services for autistic adults found that limited availability of services hinders access and negatively impacts the quality of support autistic adults receive (Nemeth et al., 2024). Two other parent interview-based studies of services for autistic adults report similar findings, and note that many existing disability programs and services are not designed for autistic people, are not appropriate to their needs, and may not be available to autistic adults without co-occurring intellectual disability (Anderson & Butt, 2018; Anderson, et al., 2018).

Findings from quantitative studies report similar issues with access to services and quality of available support. A survey of parents/caregivers of autistic adolescents and young adults found that lack of information about available services and later age of diagnosis create barriers to accessing quality support (Ishler et al., 2023). Lastly, a multi-informant (autistic adults, caregivers, professionals) survey study found that staff knowledge of autism and appropriate support for autistic people was limited at many agencies (Micai et al., 2022).

Several instruments have been developed to measure quality of support and related constructs for individuals with disabilities (Bradley & Hiersteiner, 2022; Friedman, 2018; Towson, 2012), however their functionality is limited as these tools often require a trained interviewer to administer, which is a lengthy and costly endeavor. The Quality of Care and Support (QOCS) scale (Lucas-Carrasco et al., 2011), on the other hand, is a relatively short questionnaire and was specifically designed for *and with* individuals with disabilities (Lucas-Carrasco et al., 2011). Items were initially developed with input from those with lived experience through focus groups with individuals with physical and intellectual disabilities, caregivers, and other stakeholders. The final scale includes four domains: staff quality, accessibility of care, meeting of needs, and information. The *staff quality* domain includes items related to knowledge and competence of care providers, whereas the *accessibility of care* domain focuses on accessing care (such as availability of services and cost). In contrast, the other two domains are focused on support more broadly, not necessarily within the context of professional services.

More specifically, the *meeting of needs* domain focuses on whether individuals feel they have support at home and more generally (e.g., support for leisure and activities), and the *information* domain assesses the adequacy of information an individual receives about their disability and associated services, regardless of from whom the information comes. The four-factor structure was validated separately in a sample of individuals with physical disabilities and a sample with intellectual disability.

While the QOCS has a variety of strengths, its validity for use with autistic adults in particular is unclear. Measurement tools such as questionnaires are designed to be used for a designated purpose; thus, instruments are only valid insofar as they have been tested with a *specific* population for a *specific* use (American Educational Research Association, American Psychological Association, & National Council on Measurement in Education, 2011). This means that a questionnaire that is developed for and validated in one group (e.g., people with intellectual disability) is not necessarily going to be valid and useful for another group (e.g., autistic individuals). Additionally, there could be a variety of issues related to accessibility for autistic individuals, such as the use of imprecise language that makes items difficult to understand or answer, double-barreled questions, and ableist implications (Nicolaidis et al., 2020). Recently, there has been increased attention given to validating patient-reported outcome measures (PROMs) specifically for use with autistic individuals (Nicolaidis et al., 2020; Schlitz et al., 2024). Prior research has shown that some questionnaires that have been validated in non-autistic populations do not always perform similarly well when given to autistic people, for example because autistic individuals may interpret items differently than non-autistic people, or the construct may present itself differently in autistic people (Schlitz et al., 2024).

Adapting and/or developing PROMs for use with autistic adults has been a main goal of the Academic Autism Spectrum Partnership in Research and Education (AASPIRE), a community-based participatory research (CBPR) group dedicated to conducting research that is relevant and beneficial to the autistic community from *their* perspective. Nicolaidis and colleagues (2020) detail strategies to adapt surveys based on community member input, including additional context/explanation, simplified/straightforward sentence structure and vocabulary words, links with definitions for problematic terms or clarifications, addition of graphics to response options (such as a visual analog scale with faces), and the addition of items that are specifically relevant to autism. More recently, AASPIRE conducted a “CBPR-Nested Delphi Process” to identify measurable outcomes of high priority to the autistic community (Nicolaidis et al., 2025). Community leaders, professionals, and academic experts taking part in the process selected satisfaction with / adequacy of social services and support as high-priority outcomes and identified the QOCS as a good candidate instrument for measuring this construct. Though panel members felt that the QOCS needed little adapting, they did feel the measure would be best conceptualized as two separate instruments: one to assess adequacy of *formal* disability services, and one to assess adequacy of support *in general*, regardless of receipt of formal services. Initial psychometric testing (i.e., internal reliability, test-retest reliability, and responsivity to change) indicate that both are psychometrically valid (Nicolaidis et al., in press; see all instruments at https://aaspire.org/measurement/). This paper presents a more in-depth psychometric analysis of the latter, the AASPIRE Adequacy of Support and Information Scale (AASIS).

We address the following research questions in the current study:

1. Does the factor structure of the AASIS conform to the QOCS structure found in Lucas-Carrasco et al. (2011)?
2. Is the AASIS invariant, i.e., does it perform similarly across different groups of participants?
3. What factors are related to participants’ perception of feeling adequately supported as measured by the AASIS?

## Method

This study was a multisite project. The study was approved by the Portland State University Institutional Review Board (IRB), which was the coordinating institution and managed the single IRB.

All study participants provided informed consent before starting any of the study questionnaires. When applicable, participants assented and consent was provided by legally authorized representatives (see Nicolaidis et al., 2026 for more details). Participants accessed consent forms online via a REDCap survey hosted at Oregon Health and Science University (Harris et al., 2009; Minor et al., 2019). Participants or their legally authorized representatives could review and complete consent and authorization online independently or with a support person, or over the phone or in person with a study staff member depending on each participant’s desire and needs. The study team designed consent forms for study participation (Beck et al., 2025) and data sharing (Grant et al., 2025) to maximize accessibility and understandability.

While information regarding participants and study procedure are provided below, see Nicolaidis et al. (2026) for a more extensive description of the study’s recruitment, data collection, and data integrity procedures.

### Participants

A description of the entire cohort and how they were recruited is available in Nicolaidis et al. (2026). Participants were recruited to form three subcohorts: a community subcohort, recruited online and via direct outreach to community groups and individuals; a healthcare subcohort, recruited from two university health systems; and a disability services subcohort, recruited from two regional developmental disability services systems. Recruitment differed for each subcohort – community subcohort participants were recruited via online advertisements and messages distributed via social media and email, healthcare subcohort participants were invited to participate based on review of electronic medical records, and disability services subcohort participants were recruited from local disability services agencies. Two of the study sites were explicitly designated to recruit participants into the Healthcare Subcohort (n=319), and two were designated to recruit participants into the Disability Services Subcohort (n=266). All participants were at least 18 years old and identified as being autistic (with a clinical diagnosis or self-identification), and lived in the U.S.. Participants in the healthcare and disability subcohorts had to also have a diagnosis related to the autism spectrum in their medical or service records, respectively, and had to have received care or services in one of the participating healthcare or disability services systems. Subcohorts differed from each other in several ways; for example, those in the Community Subcohort were older and in poorer health than the other two, and, compared to the Healthcare Subcohort, those in the Disability Subcohort were younger and more likely to identify as male, heterosexual, and cisgender (Nicolaidis et al., 2026).

Participants (N = 798) who completed the AASIS were included in the current investigation. Most of these participants (n = 563, 70.6%) completed the entire survey independently, about a fifth (n = 170, 21.3%) completed it with at least some support (though not necessarily on the AASIS), and 65 (8.1%) were completed entirely via caregiver report. Roughly a third of participants came from each subcohort (35.3% Healthcare Subcohort, 31.2% Disability Services Subcohort, 33.5% Community Subcohort). Participants ranged from 18-77 years old (M = 31.03, SD = 11.61) and were fairly evenly split across gender groupings (29.6% female, 38.7% male, 30.8% nonbinary/genderqueer/agender). Most (65.9%) indicated that – using their best communication method such as speech, typing, augmentative and alternative communication modalities, etc. – they could communicate fluently (i.e., discuss complicated thoughts and ideas); 20.9% indicated they could communicate by putting multiple sentences together but had trouble discussing more complicated thoughts and ideas; and 12.9% felt they could only communicate very basic ideas or feelings (such as answering yes/no, indicating hunger or pain) or communicate a small number of simple ideas or feelings (0.3% did not answer this question).

### Procedure

All participants completed an online survey hosted by [university] REDCap, though the exact procedure differed depending on into which Subcohort they were recruited. For example, Community Subcohort participants via online announcements, social media, list-serves, or word of mouth; Healthcare Subcohort participants were identified via medical records. At one of the disability services sites, study staff had access to a list of all individuals receiving developmental disability services in the county and directly contacted potential participants on this list. At the other Disability Subcohort site, study staff contacted local disability service providers in order to get connected with participants. In some cases, study staff made onsite visits to recruit participants. Across all sites, participants could take the survey independently or receive support from another person (which was sometimes a study staff member). If individuals were not able to complete the survey directly with or without as much support as they needed, caregivers could complete a caregiver version of the survey where they provided information about the individual. Participants completed surveys at three time points (baseline and follow-ups at approximately 6- and 12-months) with a random 15% subset of participants also completing an additional survey 2 weeks after the second time-point to assess test-retest reliability.

All REDCap surveys began with an informed consent form, followed by a battery of questionnaires. To reduce fatigue, the baseline survey was broken up into two parts. Participants received a $30 gift card for completing each part of the baseline survey and each follow-up survey.

### Measures *AASIS*

The AASIS is an adapted version of two subscales (*Meeting Needs*–renamed *Support* for the AASIS–and *Information*) of the QOCS (Lucas-Carrasco et al., 2011). Minor wording changes were made to the QOCS (e.g., the word “care” was removed from questions and/or replaced with “support”; e.g., “Do you feel safe about the care you get?” was updated to “Do you feel safe about the support you get?”). While the self-report version of the QOCS uses a 3-point Likert scale, the AASIS uses a 5-point Likert scale, using the same anchors as the proxy-version of the QOCS (i.e., 1=not at all, 2=a little, 3=moderately, 4=mostly, 5=totally). As recommended by community partners, an additional answer choice of “I don’t need any help” was added to the Supports subscale so as to avoid confusion for individuals who did not feel like they needed support in a given area. Thus, “I don’t need any help” was coded as a 5 (akin to being “Totally” supported), given that all of these individuals would feel satisfied with their support. The Supports subscale includes five questions assessing whether individuals get enough help with living in their home, taking part in leisure activities and social activities, and whether they get enough support and feel safe about the support they receive. The Information subscale includes four questions assessing whether participants receive enough information about their disability, services and supports, and financial benefits, as well as whether the information is easy to understand.

### Measure of Convergent Validity

#### Social Support

The AASPIRE Social Support Scale (Nicolaidis et al., in press) is an 21-item questionnaire covering five domains of support: companionship, emotional, informational, practical, and social. It is a major adaptation of the 5 PROMIS Social Support Scales corresponding to these domains (Hahn et al., 2014). This adapted measure was also tested and validated as part of the overarching study. Mean scores of all items were used in the current analyses.

### Covariate Measures for Regression Analyses

#### Demographics

We used the AASPIRE Demographic Characteristics Module (Nicolaidis et al., in press) to collect information about age, gender, sex assigned at birth, living situation, race/ethnicity, and autism diagnostic status (formal diagnosis versus self-identification). To assess communication fluency, we used the AASPIRE Communication Characteristics Module (Nicolaidis et al., in press). This question asks participants to rate their communication, under good conditions, using whatever communication modalities work best for them. Answer choices included communicating basic ideas/feelings, communicating a small number of simple ideas or feelings, communicating by putting together multiple sentences (but with trouble discussing more complicated thoughts), and communicating fluently. For the current analyses, the first two answer choices were grouped together to ensure group size allowed for comparisons. These variables were chosen based on prior literature search as well as feedback from community partners.

#### Adverse Childhood Experiences Scale

Adverse childhood events (ACES) were assessed using the ACES – AASPIRE scale (Nicolaidis et al., in press), which was a minor adaptation of the Adverse Childhood Experiences Scale (Fellitti et al., 1998). A count of the total number of ACES out of 10 was used in the current analyses.

#### Experience of Discrimination

Participants’ experience of discrimination was assessed using the Everyday Discrimination Scale - AASPIRE (Nicolaidis et al., in press), which was a minor adaptation of the Everyday Discrimination Scale (Williams et al., 1997). The scale contained 12 items rated from 0 (never) to 5 (almost every day). Questions on this scale asked about discrimination broadly (not linked to any particular characteristic or demographic group). Mean scores were used in the current analyses.

#### Health conditions

Participants’ physical and mental health were also assessed using items from the AASPIRE Health Characteristics Module (Nicolaidis et al, in press). We used a count of chronic health conditions and a dichotomous variable to indicate the presence of a mental health condition.

## Data Analysis

We conducted a confirmatory factor analysis (CFA; Brown, 2015) using weighted least squares estimation (WLSMV) to validate the two-factor structure of the AASIS. We assessed fit using standard fit indices including the root mean square error of approximation (RMSEA), the comparative fit index (CFI), the Tucker–Lewis index (TLI), and the standardized root mean square residual (SRMR). Interpretation of fit indices was guided by Hu and Bentler (1999), i.e., good fit is indicated by RMSEA of about 0.06 or less, CFI and TLI of about .95 or greater, and SRMR of about .08 or below. AASIS item reliability (overall and at the subscale level) were assessed with Cronbach’s alpha. For these analyses, all participants were included (i.e., those who directly self-reported, self-reported with support, and caregiver-reported) initially, as this is likely how the instrument would be used in a community setting. Measurement invariance testing (reported below) confirmed that combining the participants in this way is acceptable.

Correlations between AASIS total and subscale scores and scores on the AASPIRE Social Supports scale (total score and subscales) were used as an indicator of convergent validity. We hypothesized that all correlations would be positive and moderately strong, given that the AASIS and the AASPIRE Social Supports scale assess similar – yet not identical – constructs. We expected particularly high correlations between the AASIS Supports subscale and AASPIRE Social Support Scale–Practical Supports subscale and the AASIS Information subscale and the AASPIRE Social Support–Informational Support subscale.

We used the model derived from the best-fitting CFA as the basis for evaluating measurement invariance across the following variables: reporter type (self-vs caregiver-report), communication fluency (able to communicate basic ideas/feelings or a small number of simple ideas/feelings; communicate via multiple sentences but have trouble with more complicated thoughts/ideas; fluent communication), presence of intellectual disability (present; absent), race (White; BIPOC), gender (Male; Female; Nonbinary/Genderqueer/Agender).

We then used multiple logistic regression to assess variables associated with feeling adequately supported. Feeling “adequately supported” was defined as an average subscale score of at least 4.0 (corresponding to mostly or totally supported), whereas an average score of less than 4.0 was considered “not adequately supported.” Separate hierarchical models were run for the two subscales of the AASIS (Supports and Information); the first step included demographic variables only, while the second step included variables identified by the research team and community partners as relevant to feeling supported (i.e., mental and physical health, adverse childhood events, and experience of discrimination). After estimating logistic regression models for each subscale with and without the full set of variables, we calculated the predicted probability of feeling adequately supported at different levels of categorical variables that were significantly associated with feeling supported (e.g., we estimated probability of feeling adequately supported for members of each Subcohort). In these calculations, all other variables were held at their mean.

CFA and measurement invariance analyses were performed using MPlus 8.11 (1998-2017). Descriptive statistics and regression analyses were conducted with SPSS version 31.0 (IBM Corp, 2025).

## Results

### AASIS Psychometric Properties

#### Construct Validity: Confirmatory Factor Analysis

The CFA (n = 776) indicated good to excellent fit for a two-factor structure across all fit indices with the exception of the RMSEA: RMSEA = .120 [95% CI = 0.109, 0.132]; CFI = 0.969; TLI = 0.957; SRMR = 0.051. All items loaded as expected onto their respective factors (see Figure 1). The two factors had a correlation of r = .588.

**Figure 1.**
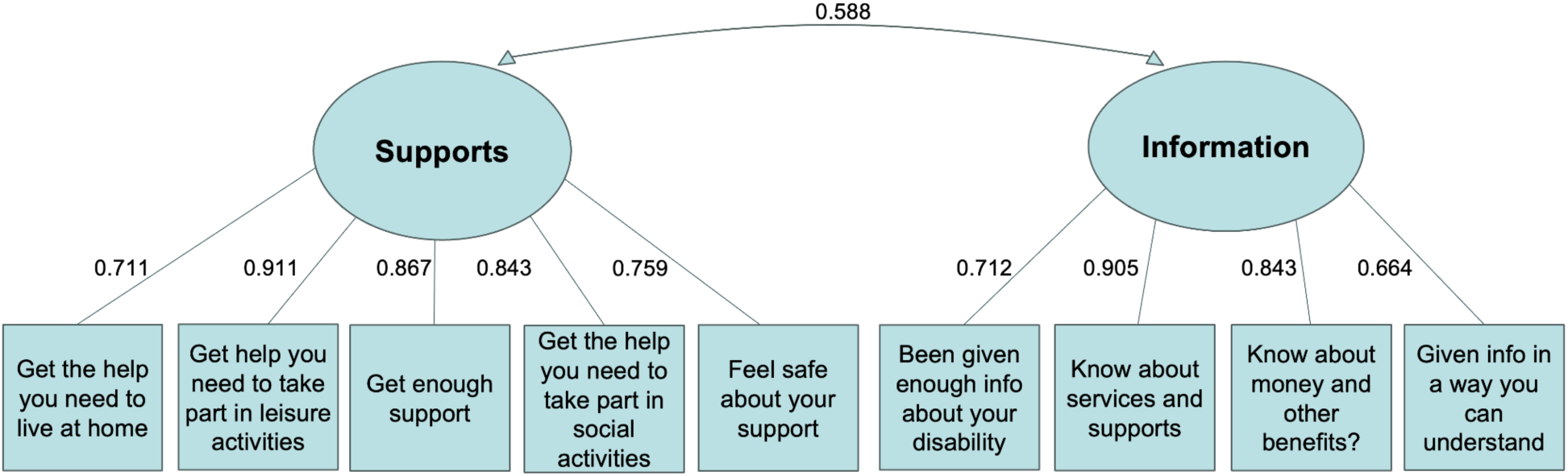
AASPIRE Adequacy of Supports and Information Scale Item Loadings.

#### Convergent Validity

The AASIS showed good convergent validity (Table 1) with the Social Support Scale. The total scores for both scales were moderately strongly correlated (r = .571), indicating that they are assessing similar yet distinct constructs.

**Table 1.** Correlations between AASIS and Subscales of the AASPIRE Social Support Scale.

|  | Total AASIS | AASIS-Supports | AASIS-Information |
| --- | --- | --- | --- |
| Companionship Support | .400 | .381 | .290 |
| Emotional Support | .491 | .437 | .395 |
| Informational Support | .488 | .450 | .375 |
| Practical Support | .390 | .353 | .306 |
| Social Isolation | .476 | .432 | .372 |
| Total Social Support | .571 | .522 | .442 |

*Note.* All correlations were significant at the $p < .001$ level. N ranged from 795 to 798.

#### Scale Reliability

The AASIS items exhibited high reliability amongst the entire scale (Cronbach’s α = .810) and both subscales (Supports: α = .864; Information: α = .801).

#### Measurement Invariance

The best-fitting CFA model showed evidence of measurement invariance of factor loadings and thresholds across relevant variables (Table 2). More specifically, the magnitude of the fit reduction for race and gender was within the suggested cut-off ranges (CFI < 01, RMSEA > .01) (Putnick & Borenstein, 2016). Although the CFI change was outside of the recommended range for the ID status and informant type, and RMSEA and CFI changes were outside of the recommended range for communication fluency, examination of the magnitude of differences in factor loadings and intercept/threshold provide evidence for invariance (i.e., the magnitude of differences in standardized factor loadings was <.15 and <.50 for standardized intercept/threshold across all the relevant models; Putnick & Borenstein, 2016).

**Table 2.**
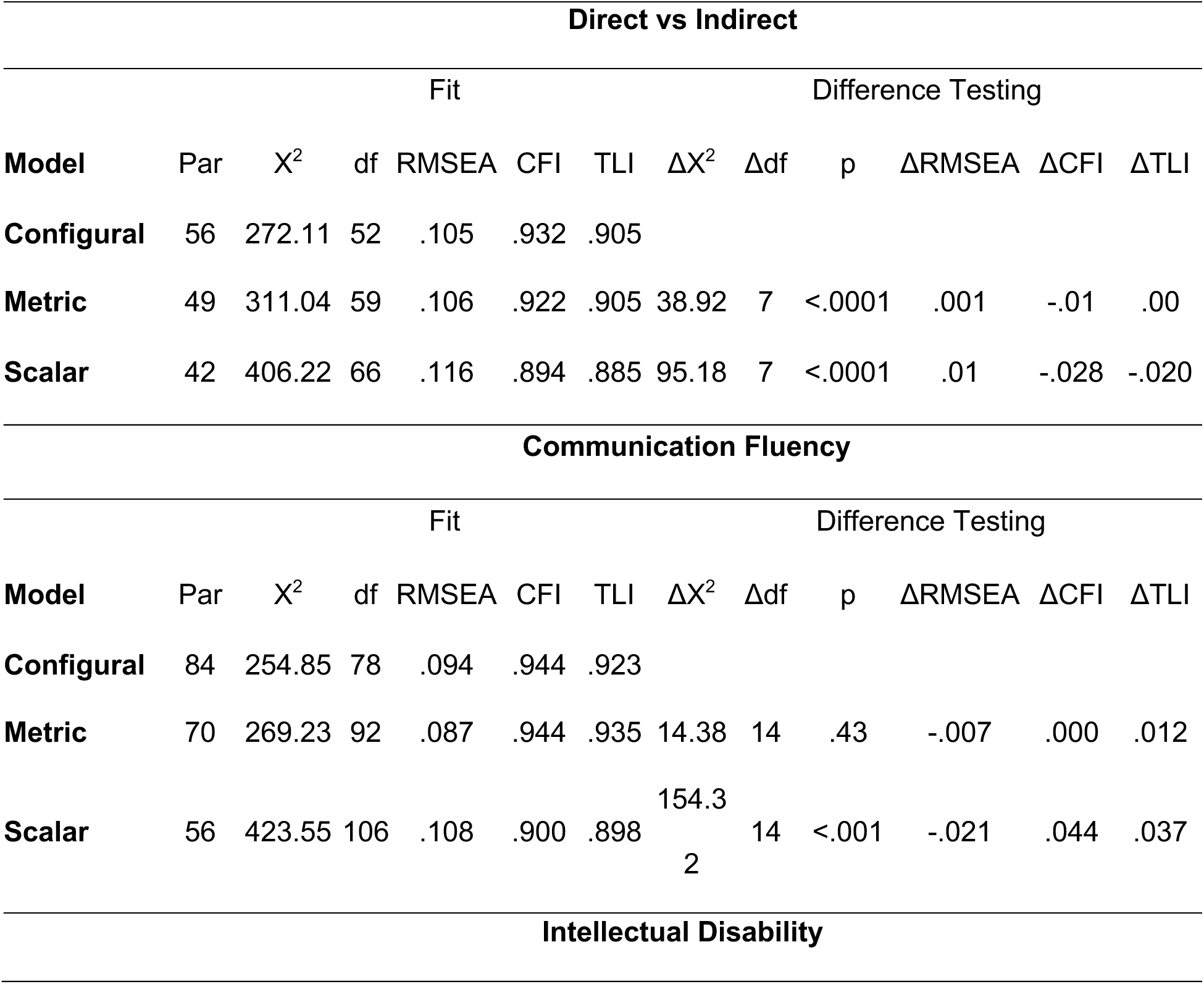
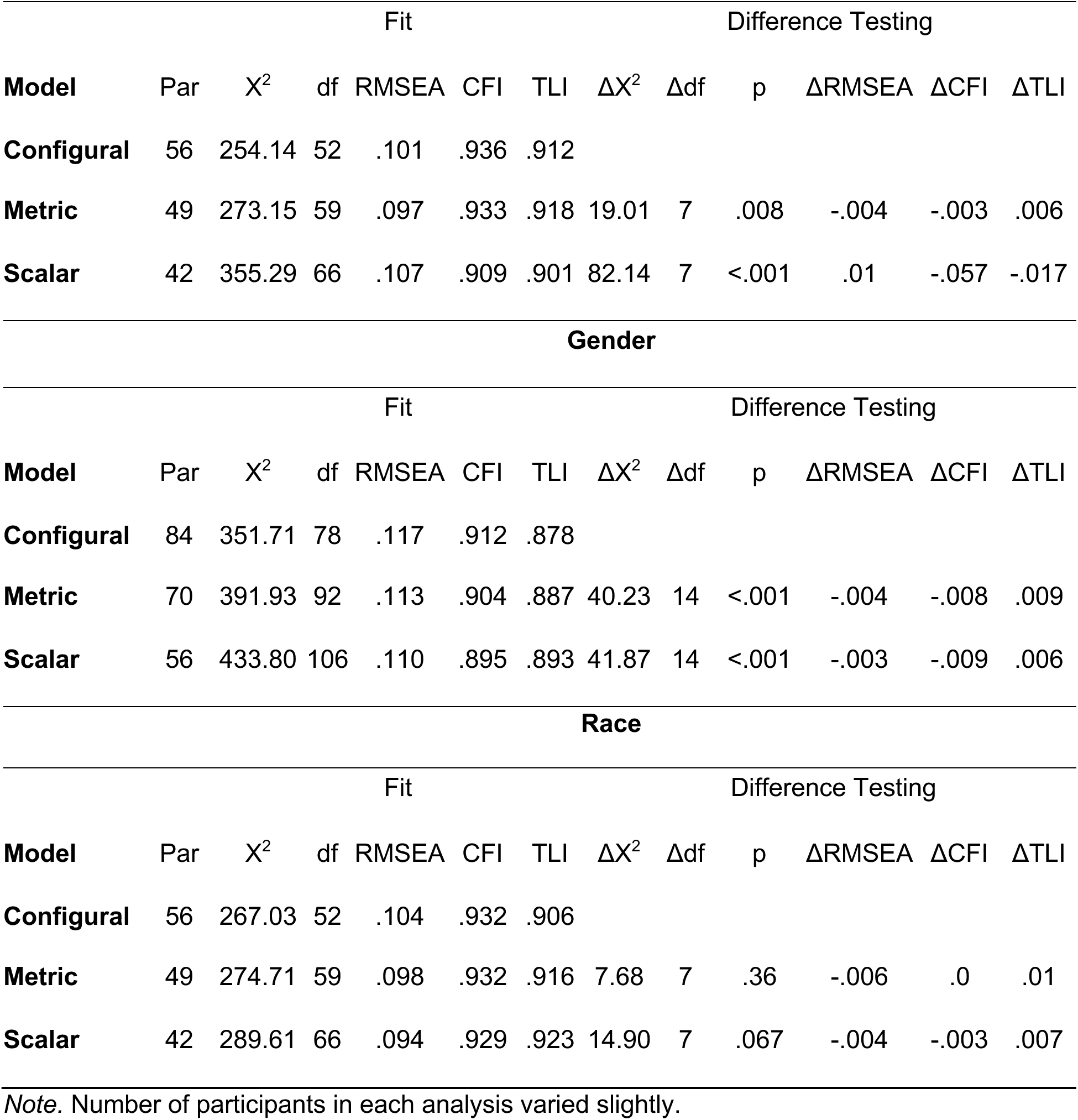
Measurement Invariance Testing.

| Direct vs Indirect |  |  |  |  |  |  |  |  |  |  |  |  |
| --- | --- | --- | --- | --- | --- | --- | --- | --- | --- | --- | --- | --- |
| Fit |  |  |  |  |  |  | Difference Testing |  |  |  |  |  |
| Model | Par | X<sup>2</sup> | df | RMSEA | CFI | TLI | ΔX<sup>2</sup> | Δdf | p | ΔRMSEA | ΔCFI | ΔTLI |
| Configural | 56 | 272.11 | 52 | .105 | .932 | .905 |  |  |  |  |  |  |
| Metric | 49 | 311.04 | 59 | .106 | .922 | .905 | 38.92 | 7 | <.0001 | .001 | -.01 | .00 |
| Scalar | 42 | 406.22 | 66 | .116 | .894 | .885 | 95.18 | 7 | <.0001 | .01 | -.028 | -.020 |
| Communication Fluency |  |  |  |  |  |  |  |  |  |  |  |  |
| Fit |  |  |  |  |  |  | Difference Testing |  |  |  |  |  |
| Model | Par | X<sup>2</sup> | df | RMSEA | CFI | TLI | ΔX<sup>2</sup> | Δdf | p | ΔRMSEA | ΔCFI | ΔTLI |
| Configural | 84 | 254.85 | 78 | .094 | .944 | .923 |  |  |  |  |  |  |
| Metric | 70 | 269.23 | 92 | .087 | .944 | .935 | 14.38 | 14 | .43 | -.007 | .000 | .012 |
| Scalar | 56 | 423.55 | 106 | .108 | .900 | .898 | 154.3<br>2 | 14 | <.001 | -.021 | .044 | .037 |
| Intellectual Disability |  |  |  |  |  |  |  |  |  |  |  |  |

|  |  |  |  | Fit |  |  | Difference Testing |  |  |  |  |  |
| --- | --- | --- | --- | --- | --- | --- | --- | --- | --- | --- | --- | --- |
| Model | Par | X<sup>2</sup> | df | RMSEA | CFI | TLI | ΔX<sup>2</sup> | Δdf | p | ΔRMSEA | ΔCFI | ΔTLI |
| **Configural** | 56 | 254.14 | 52 | .101 | .936 | .912 |  |  |  |  |  |  |
| **Metric** | 49 | 273.15 | 59 | .097 | .933 | .918 | 19.01 | 7 | .008 | -.004 | -.003 | .006 |
| **Scalar** | 42 | 355.29 | 66 | .107 | .909 | .901 | 82.14 | 7 | <.001 | .01 | -.057 | -.017 |
| **Gender** |  |  |  |  |  |  |  |  |  |  |  |  |
|  |  |  |  | Fit |  |  | Difference Testing |  |  |  |  |  |
| Model | Par | X<sup>2</sup> | df | RMSEA | CFI | TLI | ΔX<sup>2</sup> | Δdf | p | ΔRMSEA | ΔCFI | ΔTLI |
| **Configural** | 84 | 351.71 | 78 | .117 | .912 | .878 |  |  |  |  |  |  |
| **Metric** | 70 | 391.93 | 92 | .113 | .904 | .887 | 40.23 | 14 | <.001 | -.004 | -.008 | .009 |
| **Scalar** | 56 | 433.80 | 106 | .110 | .895 | .893 | 41.87 | 14 | <.001 | -.003 | -.009 | .006 |
| **Race** |  |  |  |  |  |  |  |  |  |  |  |  |
|  |  |  |  | Fit |  |  | Difference Testing |  |  |  |  |  |
| Model | Par | X<sup>2</sup> | df | RMSEA | CFI | TLI | ΔX<sup>2</sup> | Δdf | p | ΔRMSEA | ΔCFI | ΔTLI |
| **Configural** | 56 | 267.03 | 52 | .104 | .932 | .906 |  |  |  |  |  |  |
| **Metric** | 49 | 274.71 | 59 | .098 | .932 | .916 | 7.68 | 7 | .36 | -.006 | .0 | .01 |
| **Scalar** | 42 | 289.61 | 66 | .094 | .929 | .923 | 14.90 | 7 | .067 | -.004 | -.003 | .007 |

*Note.* Number of participants in each analysis varied slightly.

### Predicting Feeling Supported Based on Participants’ Characteristics

#### Supports Subscale

In the first step of the hierarchical regression using only demographic factors as independent variables, the three significant predictors of feeling adequately supported were communication fluency, gender matching sex at birth, and subcohort (Table 3).

**Table 3.**
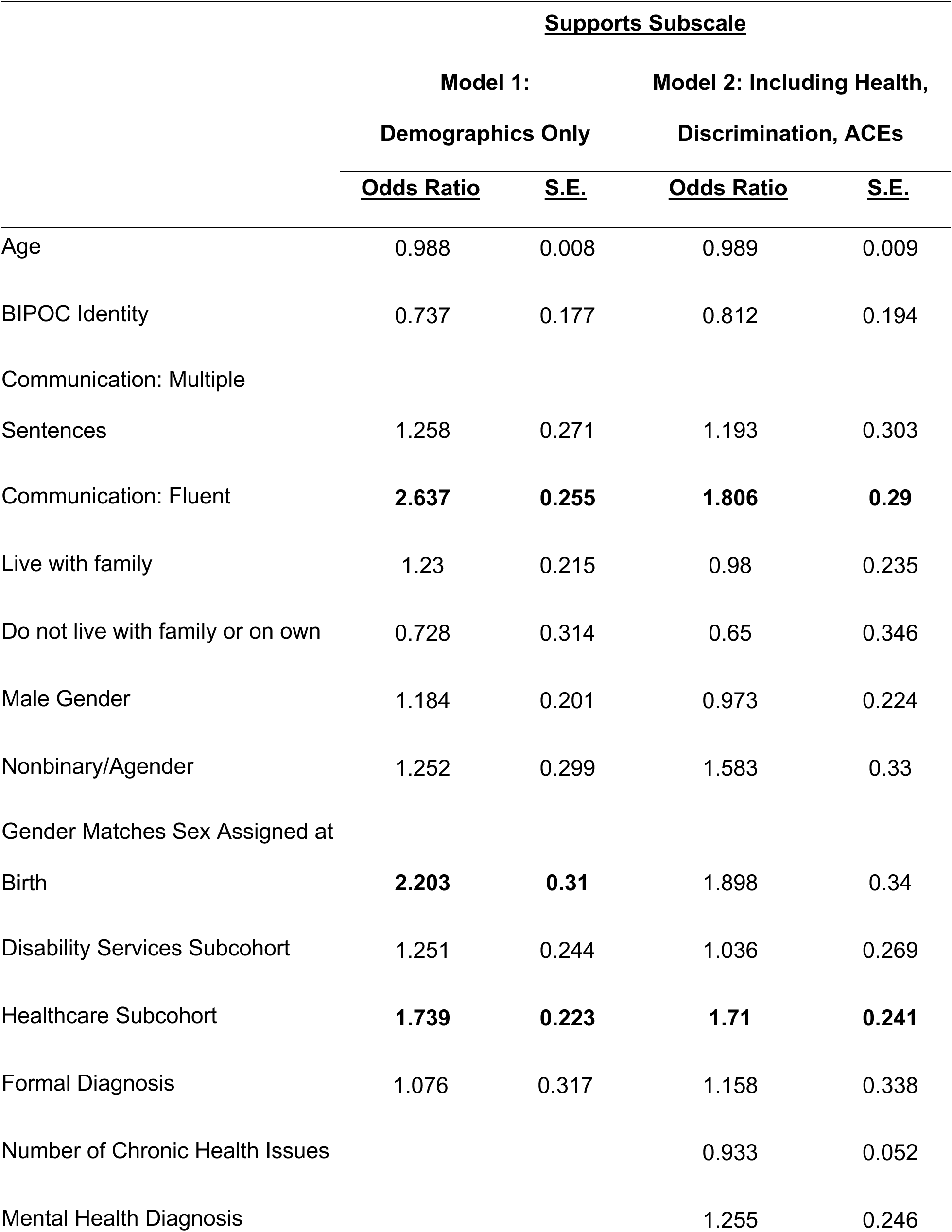
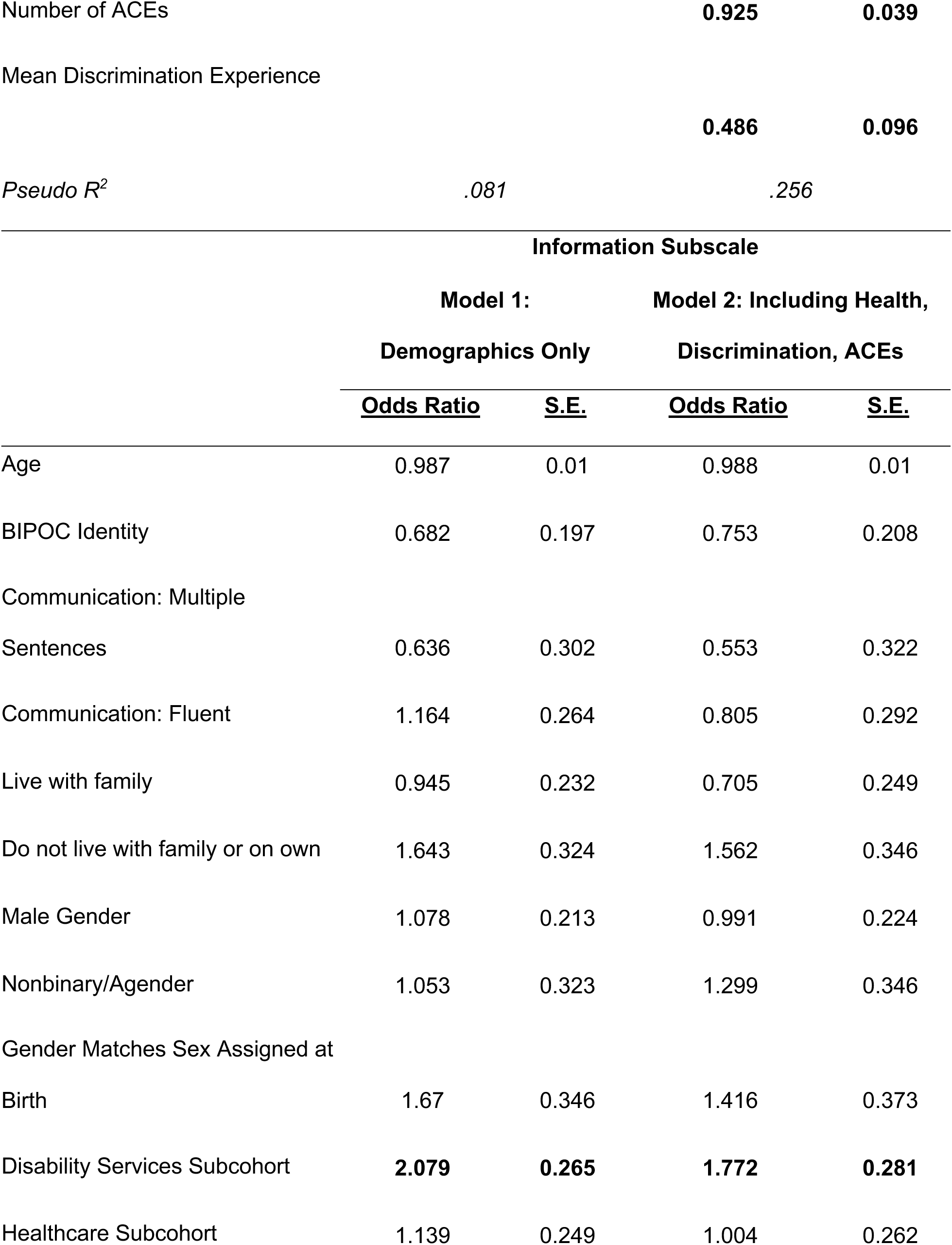
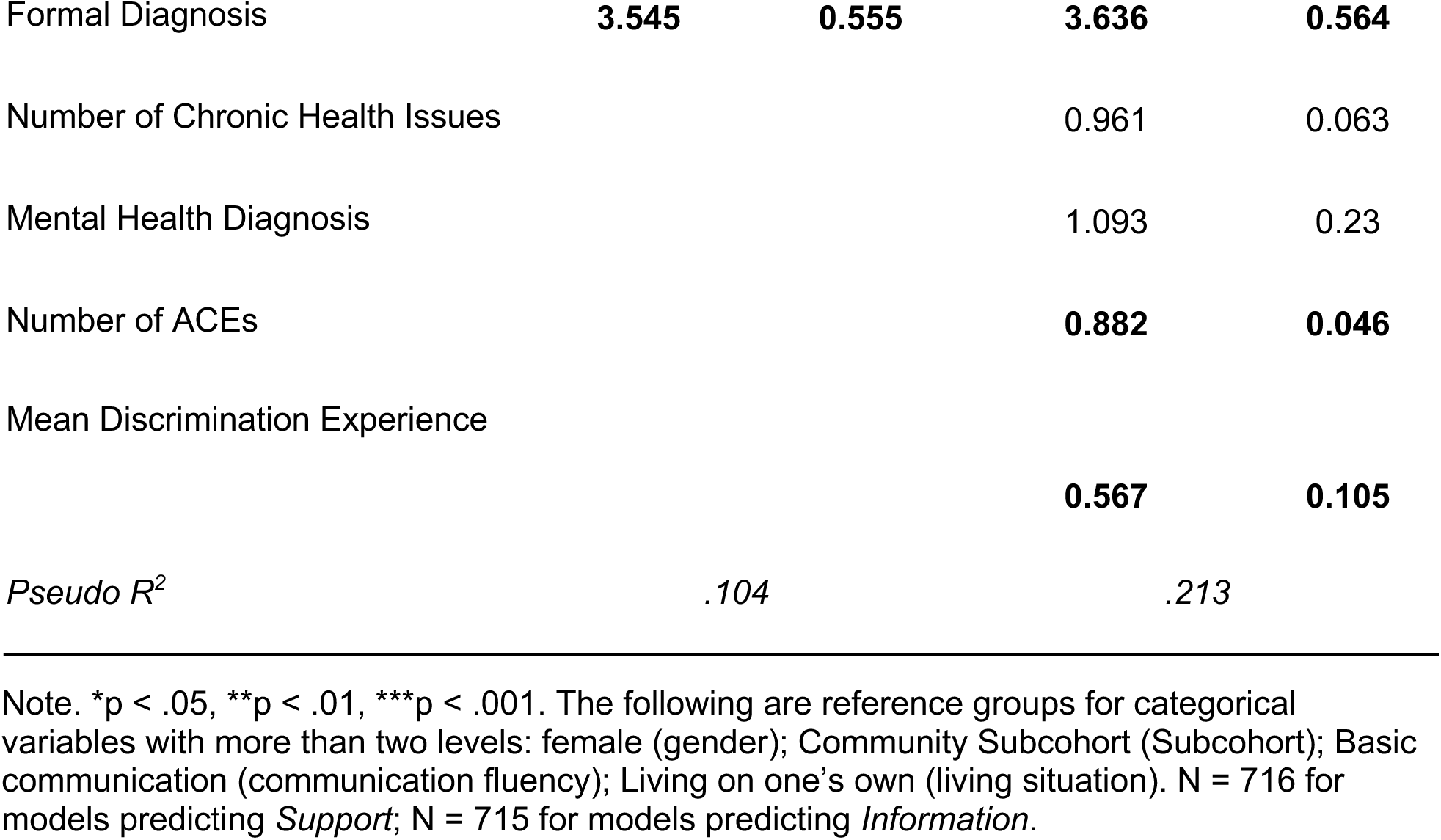
Hierarchical Logistic Regression Predicting Feeling Adequately Supported.

When physical and mental health, experiences of discrimination, and ACEs were added to the model, communication fluency and Subohort were still significant but not gender matching sex at birth Those reporting more ACES and greater experience of discrimination were significantly less likely to feel adequately supported.

Predicted probabilities of feeling adequately supported based on these subgroups are shown in Table X4a-c.

**Table 4a.** Predicted Probabilities of Feeling Adequately Supported Based on Communication Fluency.

| **Adequately Supported –**<br>**Supports Subscale** |  |
| --- | --- |
| **Demographics-Only Model** |  |
| Basic Communication | 45% |
| Multiple Sentences | 50% |
| Fluent | 68% |
| **Full Model** |  |
| Basic Communication | 52% |
| Multiple Sentences | 56% |
| Fluent | 66% |

*Note.* For each predicted probability, all other variables are held at their mean. Speech fluency was not a significant predictor of feeling supported with information in the demographics-only or full model.

**Table 4b.**
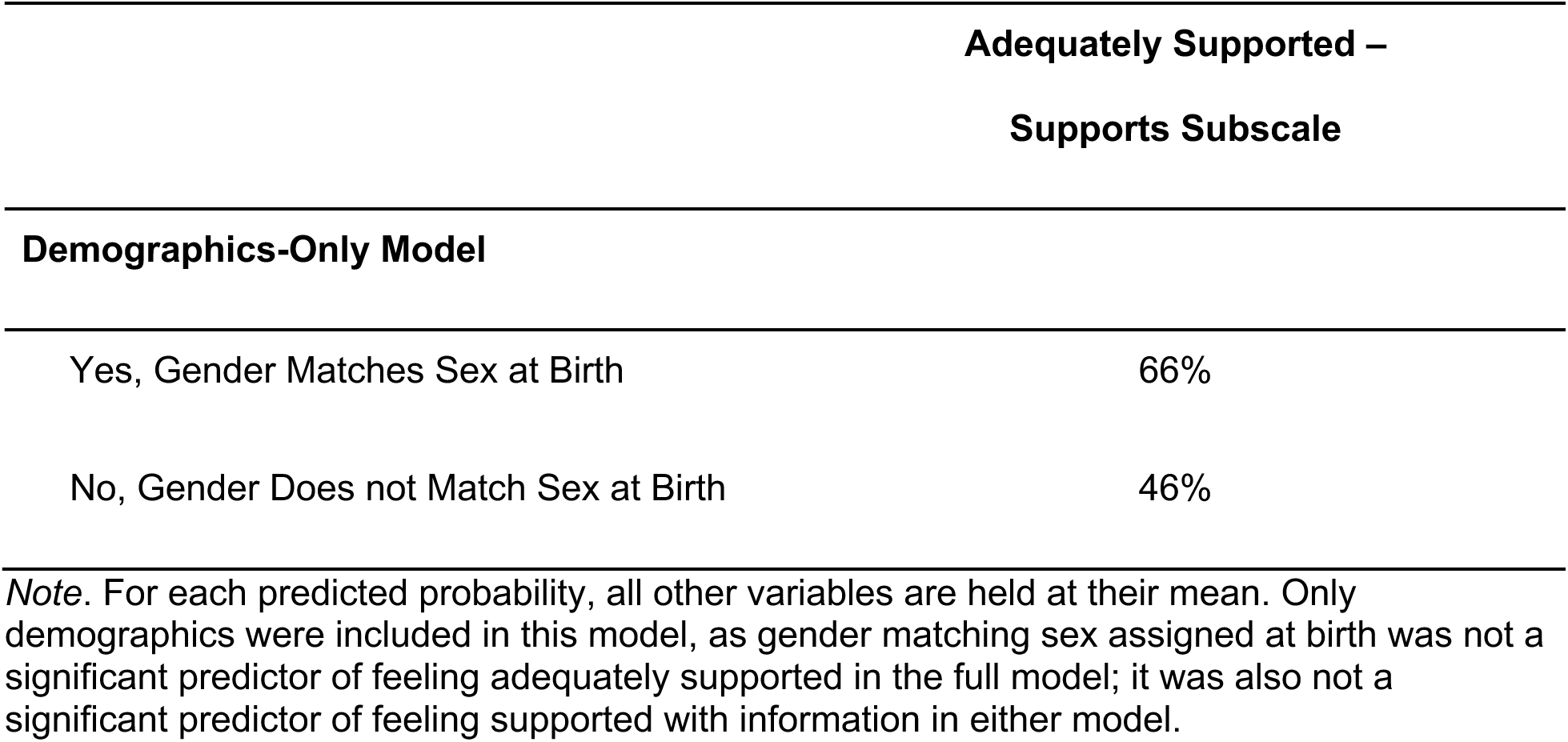
Predicted Probabilities of Feeling Adequately Supported Based on Gender Matching Sex Assigned at Birth.

**Table 4c.** Predicted Probabilities of Feeling Adequately Supported Based on Subcohort.

| **Adequately Supported –** |  |  |
| --- | --- | --- |
|  | **Supports Subscale** | **Information Subscale** |
| **Demographics-Only Model** |  |  |
| Community | 55% | 21% |
| Disability Services | 61% | 36% |
| Healthcare | 68% | 24% |
| **Full Model** |  |  |
| Community | 58% | 20% |
| Disability Services | 58% | 31% |
| Healthcare | 70% | 20% |

*Note.* For each predicted probability, all other variables are held at their mean.

**Table 4d.** Predicted Probabilities of Feeling Adequately Supported Based on Diagnostic Status.

| **Adequately Supported –**<br>**Information Subscale** |  |
| --- | --- |
| **Demographics-Only Model** |  |
| Formal Diagnosis | 29% |
| Self Diagnosis | 10% |
| **Full Model** |  |
| Formal Diagnosis | 25% |
| Self Diagnosis | 8% |

*Note.* For each predicted probability, all other variables are held at their mean. Diagnostic status was not a significant predictor of feeling supported with supports in the demographics-only or full model.

#### Information Subscale

In the first step of the hierarchical regression (including only demographic factors as independent variables), the only two significant predictors of feeling that one received adequate information about supports was Subcohort (Table 3) and having a formal, clinical diagnosis of autism.

When the other variables were added into the second step of the model, Subcohort and diagnostic status were still significant, as were the number of ACES and greater experience of discrimination, with participants with a greater number of ACES and greater discrimination being less likely to feel adequately supported in terms of receipt of information.

Predicted probabilities of feeling adequately supported based on these subgroups are shown in Table 4c.

## Discussion

Understanding disabled individuals’ perceptions of the support they receive is crucial for improving existing support and identifying areas of need that can impact overall quality of life and wellbeing. While some existing instruments designed to assess the adequacy of support for disabled people have been developed using participatory methods (Bradley & Hiersteiner, 2022; Friedman, 2018; Towson, 2012), none have specifically been developed for use with autistic people. This study further assessed the psychometric properties of the AASPIRE Adequacy of Supports and Information Scale (AASIS; Nicolaidis et al., in press), slightly adapted from the Quality of Care and Support Scale (QOCS; Lucas-Carrasco et al., 2011) using a CBPR approach. Our findings indicate preliminary validity of the AASIS and highlight potential variables that may impact autistic adults’ perception of the adequacy of support they receive.

Our CFA results indicate that the two factor solution for the AASIS (based on two of the QOCS subscales: meeting of needs and information subscales) fits relatively well in our sample of autistic adults, with fit statistics indicating good to excellent fit with only RMSEA being slightly outside of good fit range (> .1), most likely due to the sample size (Hu & Bentler, 1999), and suggesting the need for additional replication in an independent sample. Additionally, the measurement invariance analyses preliminarily support for the instrument’s invariance across several demographic factors (self-vs. caregiver report, communication fluency, intellectual disability, gender and race), suggesting that the AASIS has the capacity to function similarly across groups. This is crucial, as it tells us that individuals’ AASIS total and factor scores are comparable across people regardless of their demographic characteristics.

While further validation of the AASIS is warranted (particularly due to the high RMSEA), our regression analyses nonetheless highlight several important variables that may be related to autistic adults’ perceptions of their supports. One variable that was a significant predictor of both subscales was participant subcohort (i.e., whether participants joined the study via their healthcare provider (Healthcare subcohort), disability services provider (Disability subcohort), or through community advertisement (Community subcohort)). More specifically, holding other variables constant, compared to the Community Subcohort, those in the Healthcare Subcohort were more likely to feel satisfied with their supports (Supports Subscale) and those in the Disability Subcohort were more likely to feel satisfied with the information provided about their support (Information Subscale). With regard to feeling that support is adequate, it is possible that those in the Healthcare Subcohort represent individuals highly connected to the service system given that they or their family members had enough contact with their doctor to consent to be contacted for potential research opportunities and managed to see and follow through on messages. While it is well-documented that autistic adults face barriers receiving accessible, autism-informed healthcare (Calleja et al., 2020; Gilmore et al., 2022; Nicolaidis et al., 2015), it is possible that those who were in our Healthcare Subcohort actually represent those who have succeeded in receiving quality care and who have the most effective support to navigate a system that can be challenging and inaccessible to autistic adults. Alternatively, our findings may also reflect the fact that autistic individuals who are well connected to their healthcare system may also have highly involved, supportive families who are providing support (and thus contributing to the higher satisfaction with support).

With regard to our finding that the Disability Subcohort was the most likely to be satisfied with Information, one might wonder whether this could be explained by the number of caregiver-reports in the Disability Subcohort (11.64%), compared to only one caregiver in the Community Subcohort. However, the proportion of caregiver reports in the Healthcare Subcohort was similar to the Disability Subcohort (12.4%), yet their predicted likelihood of being satisfied with Information is still considerably less (20% in Healthcare Subcohort, 31% in Disability Subcohort). This may partially reflect the degree to which recipients of disability services engage in discussion of their services, including review of their individual goals. These discussions, which are a requirement of formal services, may provide an important source of information to autistic adults regarding the support available to them. Another potential explanation is that receipt of disability services puts autistic people in positions where their disability is frequently discussed in the context of services. For example, if an individual attends a day program which specifically serves adults with disabilities, they may be more likely to discuss their disability and services available to them with other service recipients as well as the providers who work in the program. On the other hand, this finding could reflect the fact that individuals engaged in disability services *already* have more information about their disability and support available to them. Further research is important to determine the direction of the relationship between receipt of formal services and satisfaction with information provided about support.

In both cases, the Community Subcohort were less likely to feel their support was adequate (58% adequate Supports, 20% adequate Information). While the Community Subcohort did differ in important ways from the Disability and Healthcare Subcohorts (e.g., fewer participants reporting intellectual disability, greater communication fluency, less need for help with daily living activities, etc.; Nicolaidis et al., in press), it is important to note that even when controlling for many of these or related variables, differences in predicted satisfaction by Subcohort persisted. This may be explained partly by less access to formal diagnoses and likely services, and (somewhat relatedly) higher mental health challenges, in the Community Subcohort. Furthermore, the Community Subcohort may generally be most able to mask, a phenomenon that itself may be traumatic (Cleary et al., 2023). Notably, these differences were apparent even though the regression models controlled for diagnostic status (formal or self-identified). Those who self-identified, holding all other variables stable, were less likely to feel the information they received about their services were adequate. This is likely due to the fact that self-identified autistic people are by definition not receiving services that require an autism diagnosis in order to receive insurance reimbursement. This is aligned with other research that points to self-identified autistic adults may have greater distress and unmet needs than peers with formal diagnoses (Ahuvia et al., 2026).

Greater number of ACEs and experience of discrimination were both negatively associated with feeling like Supports and Information were adequate. This could be reflective of individuals with more complex trauma histories needing more specialized, trauma-informed care than is typically available (Cobbaert et al., in press; Dodds, 2021; Peterson et al., 2019). Autistic adults with co-occurring diagnoses associated with trauma, such as Posttraumatic Stress Disorder, may feel that it is especially difficult to find information about their specific disability profile and intervention options (Quinton et al., 2024). It is also possible that some autistic adults are less satisfied in part because they experience discrimination from support providers (Cleary et al., 2023). Lastly, it is possible that those from minoritized groups – who are more likely to face discrimination – feel that they are not well understood by their support systems, leading them to feel less satisfied (Jones et al., 2025; Munday et al., 2025). This could explain why race, gender, and gender matching sex assigned at birth (in addition to number of chronic health issues or presence of psychiatric diagnosis) were not significant predictors in the full models. It is not the mere fact of belonging to one of these marginalized groups that leads to problems, but instead the discrimination and trauma that comes along with them that is harmful. The connection between gender matching sex assigned at birth and satisfaction with support in the initial regression model, using demographic factors only as predictors, is especially noteworthy given recent findings that people with disabilities who need long term services and support are more likely to be transgender than either the general population or people with disabilities who do not need LTSS, as well as a number of studies that suggest transgender identity is more common among autistic people than the general population (Kallitsounaki & Williams, 2023; Moura et al., 2025). Indeed, gender diverse autistic adults often struggle to access systems like gender-affirming healthcare and supportive relationships (Bruce, Munday, & Kapp, 2023; Munday et al., 2025). Although concordance between gender and sex assigned at birth was not significant in the full model, this may suggest broader patterns in access to and experience of support among transgender and nonbinary autistic adults which merit further research.

Lastly, communication fluency was a significant predictor of feeling support was adequate, with participants who reported fluent communication more likely to feel adequately supported than those with more basic communication abilities. This may reflect the ableism that is inherent in many service systems (McVey et al., 2023; Wang et al., 2024), and could be further influenced by the lack of access to appropriate alternative communication modalities for these participants (Donaldson et al., 2024). However, our communication fluency variable does take into account fluency in *any* modality, not just speech, so this finding may also highlight the importance of supporters truly listening to the autistic adults they support and facilitating their autonomy (Girolamo et al., 2024; Ryan et al., 2024). For instance, this could mean actively soliciting consent when providing support, and not assuming that someone is fully satisfied with the support they receive simply because they appear to comply with them.

## Limitations & Future Directions

This study provides further validation of the AASIS and identifies important factors that are associated with autistic adults’ perceptions of the adequacy of their support. However, these findings should be considered within the context of several limitations. First, this study presents data from only one survey timepoint, limiting our ability to make causal inferences. Additionally, it is difficult to collect data to adequately characterize the myriad supports our participants were potentially receiving, and there may have been aspects of the support that inform participants’ perceptions of adequacy but which are not possible for us to measure or account for in our analyses. It would also be prudent for future research on the AASIS to ensure that participants understand that, for the purpose of this instrument, “support” refers to both informal/natural and formal/paid support. While this is mentioned in the instrument instructions, it would be helpful to know whether participants thoroughly read the instructions and used that information while answering the questions. Lastly, though our sample is fairly large and is more representative of various aspects of diversity across the autism spectrum (Nicolaidis et al., 2026) than most autism studies (e.g. Maye et al., 2022; Rødgaard et al., 2022), nonspeaking individuals and those with co-occurring intellectual disability were still underrepresented. Though our invariance analyses suggest the AASIS is a valid instrument for these individuals, it is possible that there might be different predictors of satisfaction with support among these subgroups.

### Conclusion

Ensuring that the support that autistic adults receive is adequate and satisfactory is an important part of promoting health and wellbeing among this population. This is true of individuals receiving formal support (such as disability services) as well as those who are not involved in the formal service system but who nonetheless still access informal support. Using a community-based participatory approach, we collaborated with a team of autistic adults, caregivers, and relevant professionals to develop the AASPIRE Adequacy of Support and Information Scale (AASIS), a tool for measuring self-reported adequacy of support. The AASIS showed good fit into a two-factor structure, as expected, and demonstrated strong preliminary validity in our heterogeneous cohort of autistic adults. Future research with even more diverse samples (e.g., more who are nonspeaking, more with co-occurring intellectual disability), can help to further validate that the AASIS works well for these subgroups of autistic people as well as in other disability groups.

## Data Availability

All data from this study will be available through the National Institutes of Health (NIH) National Data Archive (NDA).

## Funding

This project was funded by the National Institute of Mental Health through Grant Award Number R01MH121407. It was also supported by the National Center for Advancing Translational Sciences (NCATS), National Institutes of Health, through Grant Award Number UL1TR002369. The content is solely the responsibility of the authors and does not necessarily represent the official views of the NIH.

## Acknowledgements

The authors would like to thank all the individuals who participated in this study and would also like to acknowledge the contributions of other members of the AASPIRE Outcomes Project team who made this work possible.

## Notes

### Competing Interest Statement

The authors have declared no competing interest.

### Author Declarations

The Institutional Review Board (IRB) at Portland State University (PSU) approved the project and served as the Single IRB (approval # 227674-18); Oregon Health & Science University, Vanderbilt University Medical Center, and San Diego State University waived authorization to the PSU IRB. Involvement by the Multnomah County Developmental Disability Services was subsumed under the PSU IRB.

